# Long-term outcomes of cruciate ligament injury: evidence from New Zealand linked register data

**DOI:** 10.64898/2026.08.27.26361565

**Authors:** Yana Pryymachenko, Ross Wilson, J. Haxby Abbott

## Abstract

**Objectives:** To analyse the long-term effects of a cruciate ligament (CL) injury on health and socioeconomic outcomes.

**Methods:** We used a comprehensive national injury insurance database to identify CL injuries occurring in New Zealand between 2009 and 2022, and employed a doubly robust staggered difference-indifferences research design to identify the effects of these injuries on outcomes up to 10 years after injury. The outcomes of interest were healthcare use (hospitalisations, emergency department visits, medications, knee replacement surgery for osteoarthritis), associated healthcare costs, and labour market outcomes (employment rates, income, and government benefit payments).

**Results:** We identified 61 344 CL injuries for inclusion in the analysis. Over 10-year follow-up, a CL injury resulted in increased healthcare use (0.6 more hospitalizations [95%CI 0.4 to 0.7], 1.7 more days spent in hospital [95%CI 1.3 to 2.1], 0.4 more emergency department visits [95%CI 0.3 to 0.6],2.5 more outpatient visits [95%CI 1.8 to 3.2], and 4.7 more medications dispensed [95%CI −1.8 to 11.2]) and public healthcare costs ($7 537; 95%CI 5 888 to 9 186), reduced income (−$6 060; 95%CI −11 644 to −475), and increased benefit payments ($1 152; 95%CI 542 to 1 761).

**Conclusion:** CL injuries have long-term impacts on healthcare use and socioeconomic outcomes. Strategies to reduce the incidence of CL injuries have the potential to realise large health and economic benefits.

## Introduction

Cruciate ligament (CL) injuries are serious and common conditions^1^, which incur substantial treatment costs and reduce individuals’ health-related quality of life and participation in physical activity^2,3^. As they occur predominantly in young, otherwise healthy sports participants, CL injuries have the potential for significant and long-lasting impacts on morbidity, physical activity participation, and workforce productivity and employment^4,5^.

While several studies have investigated the associations between a CL injury and development of knee osteoarthritis (OA)^6^–^8^, HRQoL outcomes, working hours and productivity^9^, no information is available on the broader long-term health and socioeconomic impacts of CL injuries. Moreover, the existing studies often suffer from narrowly defined populations that may not be generalisable, small samples with loss to follow-up, and no or inadequate accounting for confounding^9^.

New Zealand’s (NZ) unique national injury insurance scheme offers a novel opportunity to address this research gap. The Accident Compensation Corporation (ACC), a state-owned insurance provider established under the Accident Compensation Act (1972), administers a comprehensive, nationwide, no-fault personal injury insurance scheme, covering all people injured in NZ. As the sole provider of injury insurance in NZ, operating on a universal no-fault basis, the claims database of ACC therefore covers almost all CL injuries in NZ. Claims data can also be linked with nationwide, individual-level government administrative data, allowing virtually complete measurement of a broad range of health and social outcomes following injury.

The objective of this study is to exploit this unique data resource to estimate the long-term effects of CL injury on healthcare use, associated costs, and labour market outcomes.

## Methods

### Data

#### Study cohort

The study inclusion criterion was having a CL injury in NZ between 2009 and 2022 with an insurance claim accepted by ACC. As ACC provides insurance for all injuries sustained in NZ on a universal no-fault basis, these records cover almost all CL injuries occurring in NZ. We excluded people with multiple CL injuries during the study period or a previous CL injury (since the start of available ACC records in 1994), and those who were not resident in NZ throughout the entire study period (or who died before the end of the study period).

The diagnostic codes used to identify CL injuries are listed in Table A1 in Appendix A.

Injuries were classified according to the type of injury (posterior cruciate ligament tears, anterior cruciate ligament tears, sprains/strains), the activity during which the injury occurred (sports, work, other), and the sex and ethnicity (Māori [New Zealand’s indigenous population], non-Māori) of the injured individual. A small number of claims had more than one CL injury recorded; we assigned each of these to a single injury type, with the highest priority given to posterior cruciate ligament tears (i.e. a claim with a posterior cruciate ligament tear as well as other CL injuries was classified as a posterior cruciate ligament tear), followed by anterior cruciate ligament tears, and the lowest priority to sprains/ strains.

#### Outcomes

The outcomes were measured at each calendar quarter during the 2009–2022 study period. The outcomes of interest were related to healthcare use and costs (number of hospitalizations, number of days spent in hospital, emergency department visits, outpatient visits, medical prescriptions dispensed, OA-related TKR, total public healthcare costs) and labour market outcomes (income, employment, government benefit payments). Healthcare use and costs included all care received in public facilities, regardless of whether it was funded by ACC or the tax-funded public healthcare system.

All costs are reported in 2022 NZ dollars, adjusted using NZ Consumer Price Index inflation rates10 (in 2022, 1 NZD ≈ 0.64 USD).

#### Data sources

All data were obtained from the Integrated Data Infrastructure (IDI), a population-wide research database maintained by NZ’s national statistics agency, Stats NZ, linking individual-level data from multiple government administrative datasets and other sources11. Data on CL injuries were drawn from the ACC Claims dataset within the IDI. Additional linked datasets from the Ministry of Health and the Inland Revenue department were used to construct the outcome measures (see Table 1). Demographic covariates (age, sex, ethnicity) were obtained from the Personal Details dataset in the IDI, which is collated by Stats NZ from multiple administrative sources.

**Table 1:** Data sources.

| Variable | Dataset |
| --- | --- |
| **Covariates** |  |
| Age | Stats NZ, Personal Details dataset |
| Sex | Stats NZ, Personal Details dataset |
| Ethnicity (Māori, Pacific, Asian) | Stats NZ, Personal Details dataset |
| **Outcomes** |  |
| *Healthcare use and costs* |  |
| Number of hospitalisations | Ministry of Health, Publicly Funded Hospital Discharges dataset |
| Days in hospital | Ministry of Health, Publicly Funded Hospital Discharges dataset |
| Number of emergency department visits | Ministry of Health, National Non-Admitted Patient Collection |
| Number of outpatient visits | Ministry of Health, National Non-Admitted Patient Collection |
| Number of prescriptions dispensed | Ministry of Health, Pharmaceutical Claims dataset |
| Number of pain medications dispensed | Ministry of Health, Pharmaceutical Claims dataset |
| Had a total knee replacement surgery for OA | Ministry of Health, Publicly Funded Hospital Discharges dataset |
| Total public healthcare costs, NZD | Ministry of Health: Publicly Funded Hospital Discharges, National Non-Admitted Patient Collection, Pharmaceutical Claims, General Medical Subsidy Claims, Laboratory Claims datasets |
| *Socioeconomic outcomes* |  |
| *(all in NZD, except for Employment; only estimated for non-work injuries)* |  |
| Total income | Inland Revenue, Tax dataset |
| Income from wages & salary | Inland Revenue, Tax dataset |
| Employment (any wage/salary income during the period) | Inland Revenue, Tax dataset |
| Benefit payments | Inland Revenue, Tax dataset |
| ACC employment compensation payments | ACC Claims dataset |

Abbreviations: NZD = New Zealand dollars, ACC = Accident Compensation Corporation, OA = Osteoarthritis

#### Ethics and Data Availability

This study was approved by the University of Otago Human Research Ethics Committee (Health) [HD20/074]. Access to the anonymised data used in this study was provided by Stats NZ under the security and confidentiality provisions of the Statistics Act 1975. Careful consideration has been given to the privacy, security and confidentiality issues associated with using linked administrative data in the IDI; see the full disclaimer at the end of this article for further details.

The data used in this study are not publicly available due to the strict security provisions of the IDI. Access to the IDI may be made available by Stats NZ to approved researchers.

### Analytical methods

#### Identification strategy

When estimating the causal effect of a CL injury on longterm outcomes, simply comparing the outcomes of individuals who had a CL injury to those who did not may result in biased results due to unobserved characteristics (e.g. differences in physical activity level or general health status) affecting both the likelihood of receiving a CL injury and health and socioeconomic outcomes. To address this issue, we employed a quasi-experimental difference-in-differences design comparing individuals with a CL injury to individuals who had a CL later in time (and who should, therefore, have similar underlying characteristics).

The two identifying assumptions of this approach are limited anticipation and parallel trends. The limited anticipation assumption is expected to hold due to the nature of the event: a CL injury cannot be presumptively anticipated. We also expect the parallel trends assumption to hold due to the staggered event design: the control observations are from people who had a CL injury later, who can be expected to be similar in activity profiles and health status to those whose injury happened to occur earlier. We assessed the plausibility of this assumption by estimating the ‘effects’ for preinjury periods (which are expected to be near zero and constant over time).

We estimate per-period effects (for the quarter of injury, the following quarter, etc.) using a doubly-robust staggered difference-in-difference estimation method^12^. The doubly-robust estimator combines outcome regression and inverse probability weighting of exposure, and is robust to misspecification in either one of the outcome or exposure models^13^. All analyses were adjusted for age (at the end of the study period, and specified as discrete 5-year age groups to allow for non-linear associations between age and the outcome measures) to account for expected differences in the trajectory of outcomes over the life course.

To describe the overall long-term effects, we aggregated the per-period results to 5 and 10-year cumulative effects.

Effects on the labour market outcomes (income, employment, benefit payments) were estimated for non-work injuries only. It is not possible, with our analytical design, to obtain unbiased estimates of effects of injuries occurring at work on employment-related outcomes (by definition, the treated observations were employed at the start of their treatment period, so their employment rates can only go down during the follow-up; conversely, the control observations were, again by definition, employed at some point during the follow-up period of the treated observations, so their employment rates cannot go down during the follow-up period).

#### Robustness checks

To check whether the results were robust to model specification, we performed the analysis with additional controls (sex and ethnicity). We also conducted analyses for the smaller cohort of those having injuries between 2017 and 2022; this addresses both potential selection bias due to the residency and mortality restrictions (as with a shorter study period the restrictions are less stringent) and possible violations of the parallel trends assumption due to inherent differences in outcome trajectories between groups injured further apart in time.

#### Subgroup analyses

We conducted separate analyses for those with anterior cruciate ligament tears, posterior cruciate ligament tears, and CL sprains, and for those who were injured at work, during sports, or in other activities. To investigate potential inequities in the impacts of CL injuries, we also conducted subgroup analyses by ethnicity (Māori and non-Māori) and sex (male and female).

#### Software

All analyses were conducted in R (version 4.5.1)^14^, using the {did} package for doubly-robust estimation of the staggered difference-in-differences model^15^.

## Results

### Data summary

There were 116 208 CL injuries between 2009 and 2022 recorded in the ACC claims data, in 106 422 peo-ple. After excluding people with multiple or previous CL injuries (n=12 738), those who died before 2022 (n=2 520), and those not residing in NZ throughout the study period (n=29 820), the final analysis cohort consisted of 61 344 individuals and 3 435 264 quarterly observations.

Slightly more than half of the cohort were male (n=33 504, 55%) and 20% (n=12 246) were Māori. Sprains and strains were the most common type of injury (n=44 256, 72%), followed by ACL tears (n=15 999, 26%) and PCL tears (n=1 083, 2%). Most injuries were sustained during sports (n=37 830, 62%), 11% at work (n=6 828), and 27% in other or unspecified activities (n=16 689). The average age at the time of injury was 38 years. Descriptive statistics of the outcomes before and after the CL injury are available in Table A2.

#### Pre-injury trends

The estimated ‘effects’ in all pre-injury periods were close to zero for all outcomes (Figure 1), providing support for the plausibility of the parallel trends assumption.

**Figure 1:**
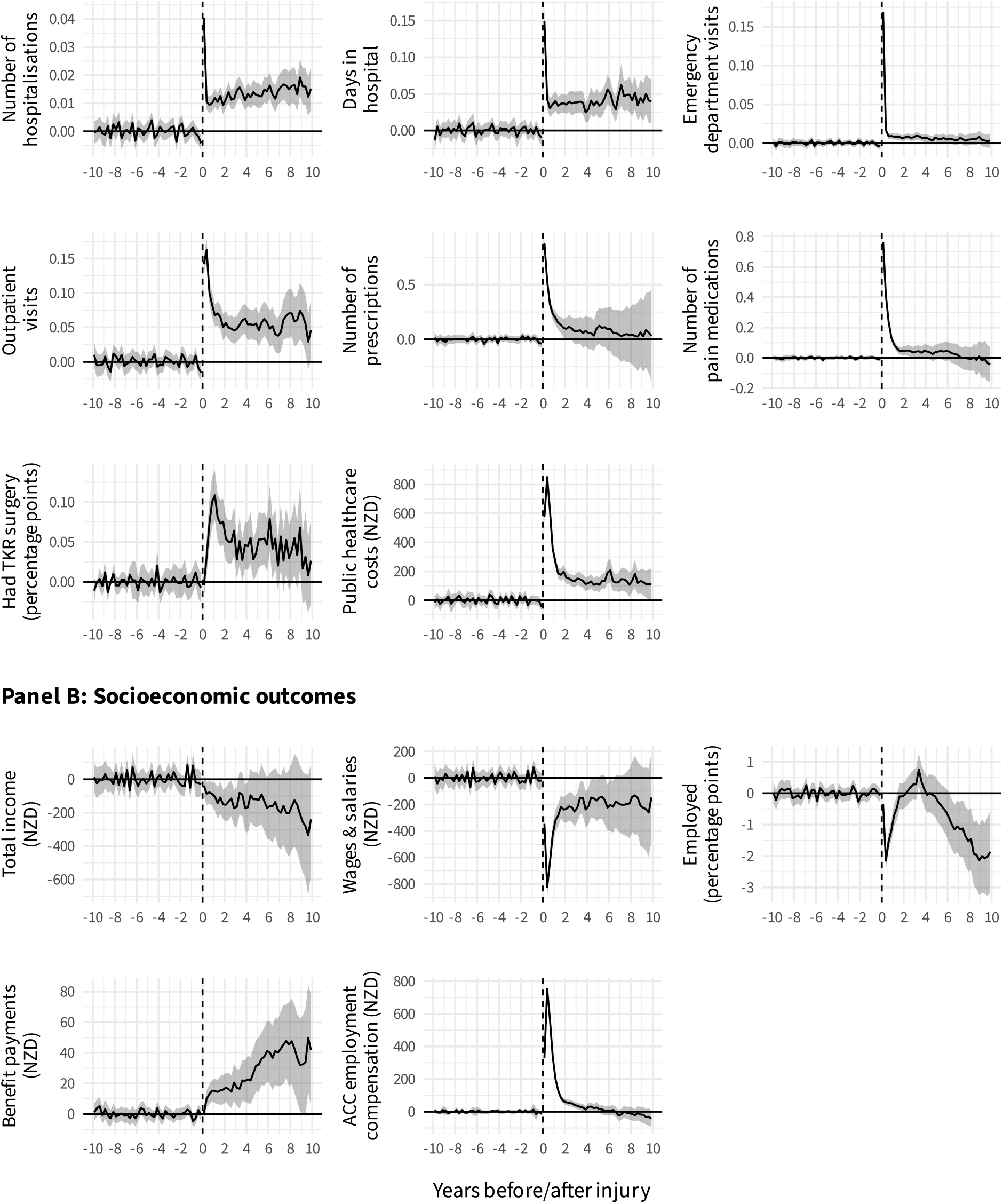
Average per-quarter effects of a cruciate ligament injury.

### Effects of CL injury

#### Healthcare use and costs and injury incidence

All of the healthcare use outcomes were increased in the quarter of the injury (Figure 1, Panel A). The rate of hospital admissions remained elevated throughout the follow-up period, with a cumulative effect of an additional 0.6 hospitalisations (95%CI 0.4 to 0.7; a 34% increase compared to the no-injury control) and 1.7 more days in hospital (95%CI 1.3 to 2.1; 73%) over 10-year follow-up (Table 2, column (1)). The effect on ED visits dropped substantially after the quarter of injury, but remained positive throughout the follow-up period, resulting in 0.4 more visits (95%CI 0.3 to 0.6; 15%) over 10 years. The effect on outpatient visits persisted throughout the follow-up period, with a cumulative 10-year effect of an additional 2.5 visits (95%CI 1.8 to 3.2; 24%). Effects on medication prescriptions declined gradually over the first 2 years following injury, with a cumulative effect of 4.7 more prescriptions (95%CI −1.8 to 11.2; 3%), including 2.5 more pain medication prescriptions (95%CI 0.6 to 4.4; 8%), over 10 years.

**Table 2:** Aggregated effects of a CL injury on long-term outcomes.

| Outcome / Aggregation period | (1)<br>Main results | (2)<br>With additional controls | (3)<br>Smaller cohort |
| --- | --- | --- | --- |
| **Healthcare use and costs** |  |  |  |
| *Number of hospitalisations* |  |  |  |
| 10 years | 0.57 (0.45 to 0.69) | 0.56 (0.44 to 0.68) |  |
| 5 years | 0.27 (0.22 to 0.32) | 0.27 (0.22 to 0.32) | 0.30 (0.20 to 0.40) |
| *Days in hospital* |  |  |  |
| 10 years | 1.73 (1.31 to 2.14) | 1.70 (1.29 to 2.11) |  |
| 5 years | 0.84 (0.65 to 1.03) | 0.84 (0.65 to 1.03) | 0.86 (0.36 to 1.36) |
| *Emergency department visits* |  |  |  |
| 10 years | 0.42 (0.26 to 0.58) | 0.43 (0.27 to 0.58) |  |
| 5 years | 0.32 (0.26 to 0.38) | 0.32 (0.26 to 0.38) | 0.29 (0.17 to 0.40) |
| *Outpatient visits* |  |  |  |
| 10 years | 2.49 (1.81 to 3.16) | 2.45 (1.80 to 3.11) |  |
| 5 years | 1.38 (1.13 to 1.63) | 1.38 (1.12 to 1.64) | 1.17 (0.71 to 1.64) |
| *Number of prescriptions* |  |  |  |
| 10 years | 4.69 (– 1.84 to 11.21) | 4.90 (– 2.32 to 12.12) |  |
| 5 years | 3.47 (1.69 to 5.26) | 3.50 (1.64 to 5.37) | 3.22 (– 1.84 to 8.27) |
| *Number of pain medications* |  |  |  |
| 10 years | 2.47 (0.58 to 4.37) | 2.65 (0.80 to 4.50) |  |
| 5 years | 2.28 (1.75 to 2.82) | 2.32 (1.74 to 2.89) | 3.27 (1.63 to 4.90) |
| *Had TKR surgery (percentage points)* |  |  |  |
| 10 years | 1.9 (1.3 to 2.6) | 1.9 (1.3 to 2.5) |  |
| 5 years | 1.1 (0.8 to 1.3) | 1.1 (0.8 to 1.3) | 1.0 (0.5 to 1.5) |
| *Public healthcare costs (NZD)* |  |  |  |
| 10 years | 7 537 (5 888 to 9 186) | 7 449 (5 608 to 9 291) |  |
| 5 years | 4 821 (4 255 to 5 386) | 4 809 (4 250 to 5 368) | 5 629 (4 045 to 7 214) |
| **Socioeconomic outcomes (only for non-work injuries)** |  |  |  |
| (all NZD, except for Employed) |  |  |  |
| *Total income* |  |  |  |
| 10 years | – 6 060 (– 11 644 to – 475) | – 6 416 (– 12 139 to – 693) |  |
| 5 years | – 2 379 (– 4 115 to – 643) | – 2 456 (– 4 295 to – 617) | – 4 334 (– 7 791 to – 876) |
| *Wages & salaries* |  |  |  |
| 10 years | – 9 436 (– 14 675 to – 4 198) | – 9 752 (– 15 516 to – 3 987) |  |
| 5 years | – 5 669 (– 7 234 to – 4 104) | – 5 765 (– 7 637 to – 3 892) | – 8 923 (– 12 539 to – 5 307) |
| *Employed (quarters)* |  |  |  |
| 10 years | – 0.31 (– 0.52 to – 0.10) | – 0.30 (– 0.51 to – 0.08) |  |
| 5 years | – 0.05 (– 0.12 to 0.03) | – 0.05 (– 0.12 to 0.02) | – 0.11 (– 0.26 to 0.04) |
| *Benefit payments* |  |  |  |
| 10 years | 1 152 (542 to 1 761) | 1 122 (521 to 1 723) |  |
| 5 years | 349 (164 to 533) | 348 (176 to 521) | 354 (– 29 to 737) |
| *ACC employment compensation* |  |  |  |
| 10 years | 2 702 (1 861 to 3 544) | 2 600 (1 744 to 3 455) |  |
| 5 years | 2 892 (2 599 to 3 186) | 2 881 (2 582 to 3 180) | 4 556 (3 940 to 5 171) |

95% confidence intervals in parentheses. Column (1) controls for age (using 5-year age group dummy variables). Column (2) additionally controls for sex and Māori and Pacific ethnicities. Column (3) has the same controls as column (1).

The rate of TKR for OA was also increased after CL injury, although the magnitude of the effect was small in absolute terms: a cumulative effect of 0.019 (95%CI 0.013 to 0.026; 137%), or one additional surgery for every 52 people injured. The overall effect on healthcare costs was an increase of $7 537 (95%CI $5 888 to $9 186; 41%) over the 10 years following injury.

#### Socioeconomic outcomes

CL injury resulted in reductions in wages, employment rates, and total income, and increased government benefit payments, throughout the 10-year follow-up period (Figure 1, Panel B). Aggregated over the followup period, individuals with a non-work CL injury received $1 152 more government benefit payments (95%CI $542 to $1 761; 9%), $9 436 less wages (95%CI −$14 675 to −$4 198; −2%) and $6 060 less total income (95%CI −$11 644 to −$475; −1%) compared to the no-injury control (Table 2, column (1)).

#### Robustness checks

The results were robust to inclusion of additional controls (Table 2, column (2)). Restricting the sample to CL injuries between 2017 and 2022 yielded similar effects to the primary analysis (over 5-year horizon) on healthcare use outcomes but slightly stronger effects on socioeconomic outcomes (Table 2, column (3)).

#### Subgroup analyses

The effects on all outcomes except for TKR were larger for PCL tears compared to ACL tears, and for ACL tears compared to sprains (Figure 2, upper panel of each subfigure). There were no consistent differences in the effects on healthcare use and costs for injuries occurring during sports, work, or other activities, or in the socioeconomic impacts for injuries occurring during sports or other activities (Figure 2, lower panel of each subfigure). The trajectory of effects over the follow-up period by injury type and cause are presented in Figure A1 and Figure A2.

**Figure 2:**
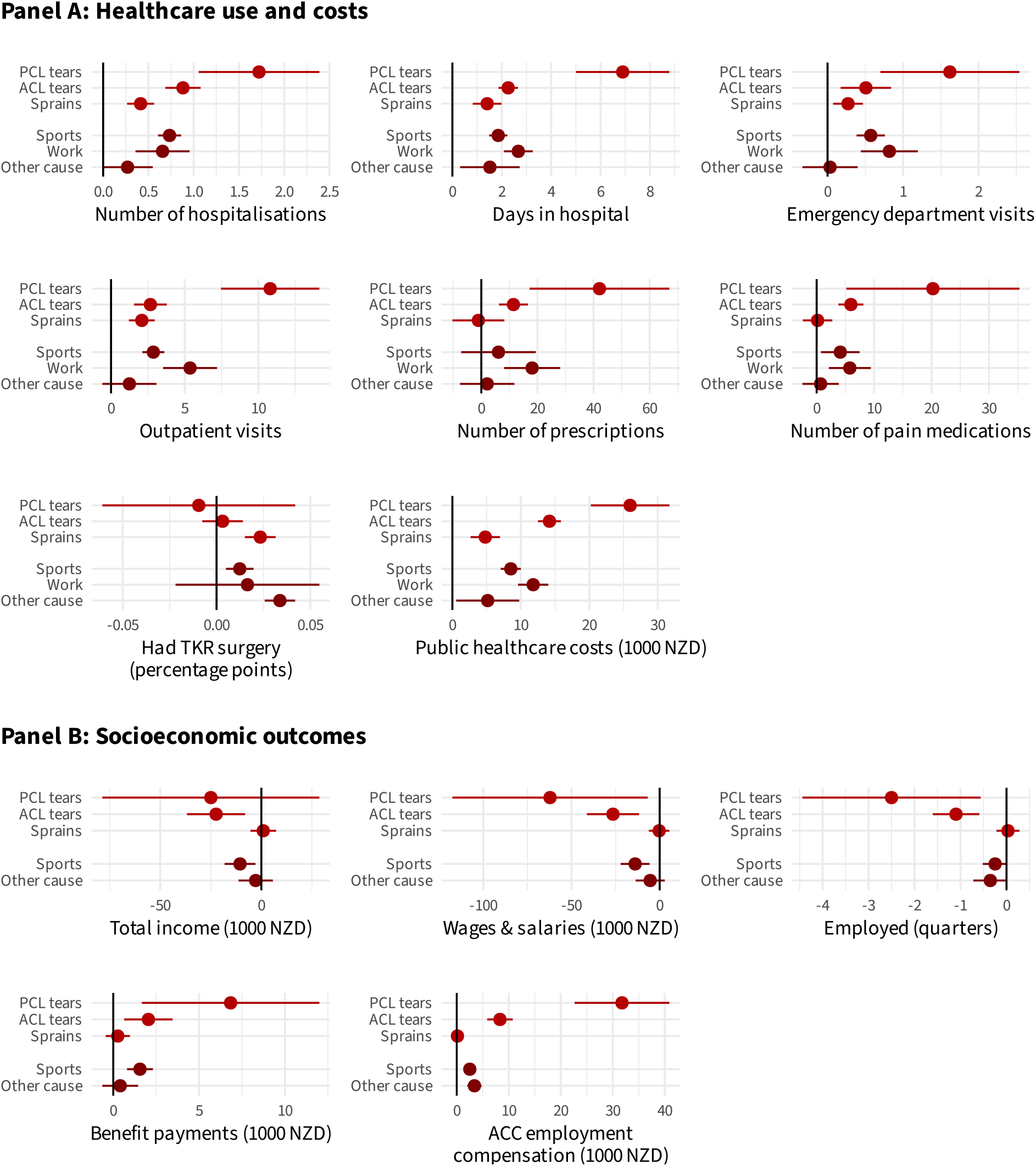
Aggregated effects of CL injury on long-term outcomes, by injury subgroups.

There was little evidence of differences in impacts of injury between population subgroups (by sex or ethnicity) for healthcare use, except for a larger effect on hospital admissions for females than for males (Figure 3) and a smaller effect on TKR for Māori than non-Māori The effects on socioeconomic outcomes were larger for males (compared to females); effects on employment and benefit payments, but not wages or income, were larger for Māori (compared to non-Māori). The trajectory of effects over the follow-up period by population subgroups are presented in Figure A3 & Figure A4.

**Figure 3:**
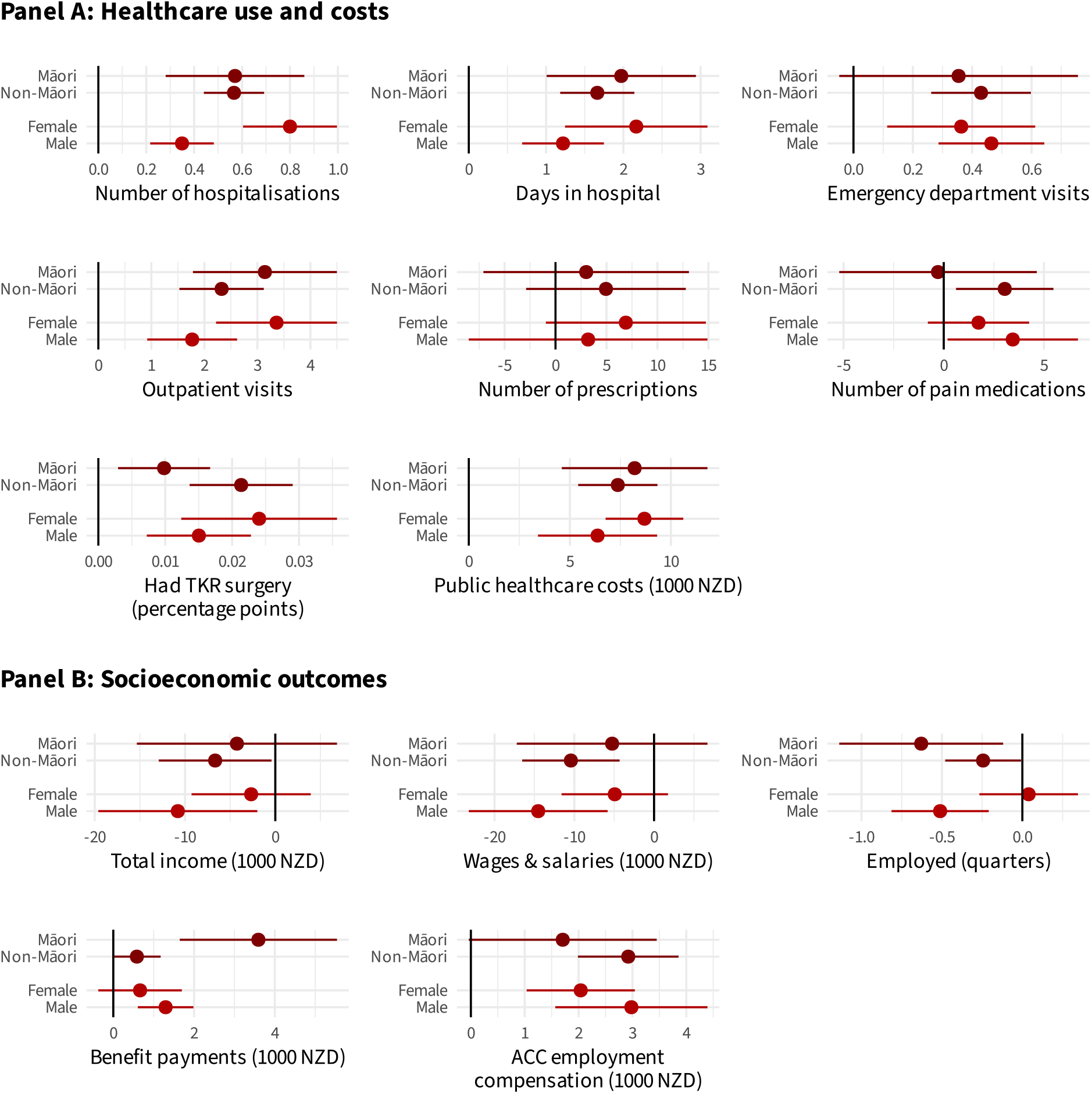
Aggregated effects of CL injury on long-term outcomes, by population subgroups.

## Discussion

This study investigated the long-term impacts of CL injury on health and socioeconomic outcomes in NZ by applying a staggered difference-in-differences approach. We found significant and long-lasting effects, including increase in healthcare utilization and costs, reduction in income and employment, increase in benefit payments, and increase in TKR associated with OA.

While there was little difference between population subgroups in the effect of CL injury on healthcare use, the effects on socioeconomic outcomes were generally larger for Māori than for non-Māori and for men than for women. These differences may reflect differences in occupational type, with Māori and men more likely to be employed in physically demanding roles that are directly impacted by the physical limitations of a CL injury. While the impact on employment was larger for Māori than for non-Māori, there was no evidence of differences in the effect on either wages or total income; this is likely due to the lower average incomes received by Māori, and highlights the need to consider measures beyond direct income loss when evaluating the socioeconomic impact of injury.

### Strengths and limitations

A key strength of this analysis is the use of a staggered difference-in-differences design to enable causal identification of the effects of CL injuries. Under plausible identifying assumptions, our results admit a causal interpretation without confounding by pre-injury status. In previous analyses of these data, we have identified that comparison of injured and uninjured samples, even after matching or adjustment for a large set of preinjury characteristics, is unlikely to produce unbiased estimates of the effect of injuries^16^. The approach used in this study, we argue, successfully circumvents this issue.

One potential drawback of our analysis is that the control and treatment groups are less likely to be similar the further apart in time the injuries are. When restricting the analysis to injuries no more than 5 years apart, the estimated effects (over a 5-year horizon) on healthcare use were similar to those for the full cohort, but the effects on socioeconomic outcomes were slightly larger, suggesting that our primary 10-year results for socioeconomic impacts may be underestimated. Another possible limitation is that the control observations may be selected for better outcomes; the absence of a CL injury in otherwise comparable individuals may in some cases be due to non-CL injuries or other adverse health events resulting in reduced physical activity, and thus reduced risk of CL injury. If the selected control observations disproportionately exclude people having other health events, our results are likely to overestimate the true effect of CL injuries.

Another key strength is the use of NZ’s comprehensive and detailed linked administrative data, allowing the analysis of a wide range of healthcare and socioeconomic outcomes over long-term follow-up for the entire population of interest. The universal no-fault coverage of the ACC scheme means that almost all CL injuries are captured in our data, so our results are representative of the NZ population. However, our results may have less generalisability to other healthcare settings in which comprehensive injury-related healthcare services, as well as additional support such as vocational rehabilitation and earnings compensation, that are provided by ACC are not available or are available only to some patients.

### Previous evidence

This is the first study to investigate the breadth of health and socioeconomic outcomes over long-term follow-up after CL injury. Nevertheless, our results are consistent with other evidence reported previously, which may also shed light on the mechanisms behind the long-term effects found in this study. A high share of individuals do not return to sport after an ACL reconstruction surgery^17^, and ACL injury is associated with increased work absenteeism and reduced work intensity^9^. The long-term trajectories of reduced employment and income found in this study similarly suggest that CL injuries can result in prolonged reduction in work capacity.

Knee injuries are also strongly associated with development of OA^7^,^8^,^18^. We were unable to identify OA symptom onset, diagnoses, or treatment prior to surgical intervention in our administrative data, but found an increased rate of TKR (with an OA diagnosis) after CL injury, although the absolute rate of TKR was low (an increase of less than 2 percentage points receiving surgery over 10 years).

Previous evidence has also demonstrated poorer longterm health-related quality of life^19^ after ACL injury and a high rate of re-injury following ACL reconstruction^20^. We were unable to analyse these outcomes in our study, as patient-reported outcomes were not available in the administrative datasets and our analytical design restricted the sample to people having a single CL injury.

### Implications for policy and practice

The results of this study show that CL injuries have significant and long-lasting effects on health and socioeconomic outcomes. In light of these results and considering the high prevalence of CL injuries, the implementation of injury prevention programs for groups at risk of CL injury is likely to yield considerable benefits at an individual and societal level.

## IDI Disclaimer

These results are not official statistics. They have been created for research purposes from the Integrated Data Infrastructure (IDI), which is carefully managed by Stats NZ. For more information about the IDI, please visit https://www.stats.govt.nz/integrated-data/.

The results are based in part on tax data supplied by Inland Revenue to Stats NZ under the Tax Administration Act 1994 for statistical purposes. Any discussion of data limitations or weaknesses is in the context of using the IDI for statistical purposes, and is not related to the data’s ability to support Inland Revenue’s core operational requirements.

## Appendix A. Supplementary Material

**Table A1:**
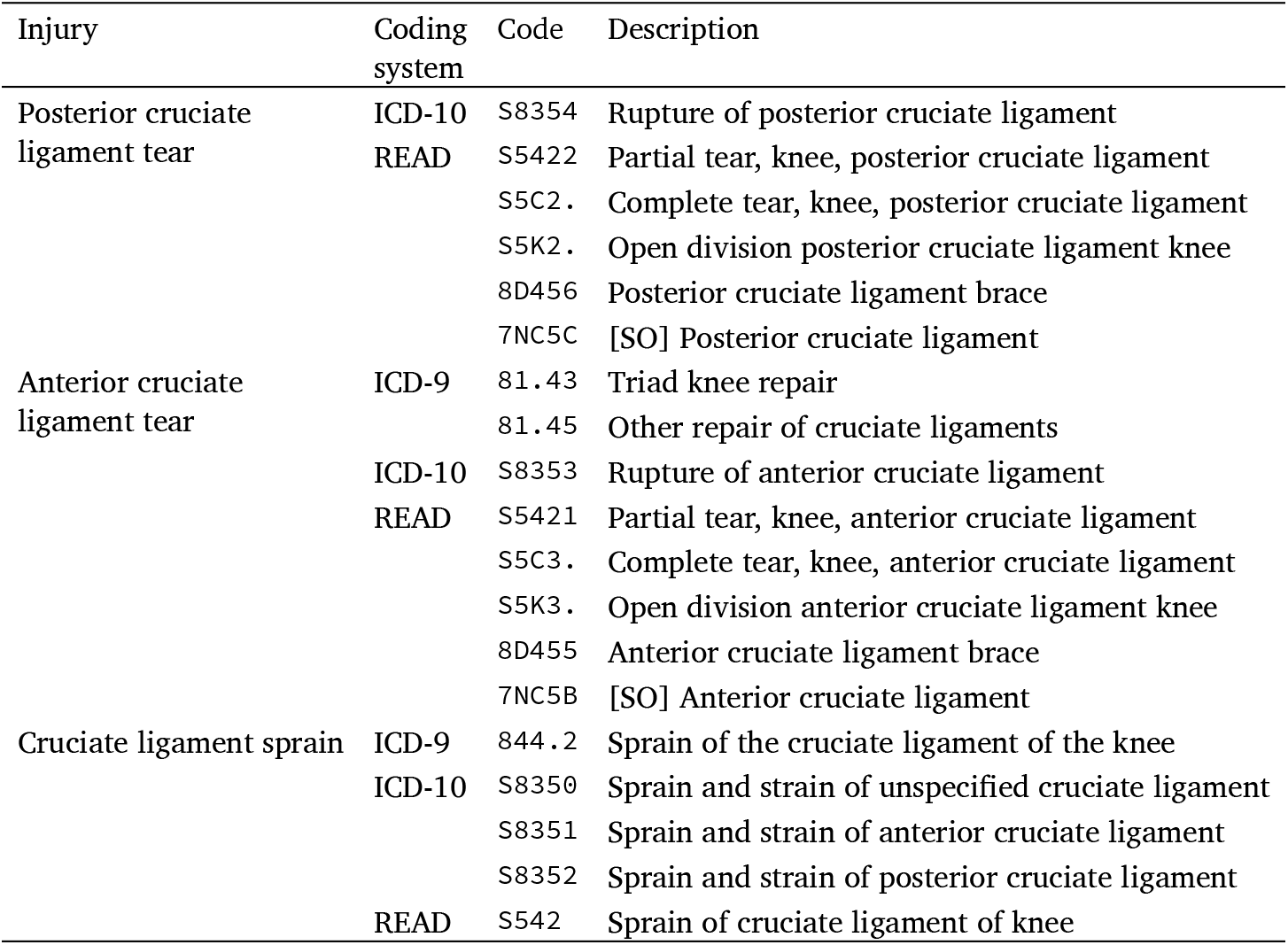
Diagnosis codes used to identify knee injuries.

**Table A2:** Descriptive statistics of the outcomes.

| Outcome | Before CL injury | After CL injury |
| --- | --- | --- |
| *Healthcare use and costs* |  |  |
| Number of hospitalisations | 0.04 (0.26) | 0.06 (0.31) |
| Days in hospital | 0.06 (0.84) | 0.10 (1.33) |
| Emergency department visits | 0.06 (0.33) | 0.08 (0.37) |
| Outpatient visits | 0.21 (1.22) | 0.32 (1.75) |
| Number of prescriptions | 2.52 (8.11) | 3.99 (10.31) |
| Number of pain medications | 0.50 (3.15) | 0.81 (3.37) |
| Had TKR surgery (N) | 240 | 1248 |
| Public healthcare costs (NZD) | 369 (2 865) | 661 (4 308) |
| *Socioeconomic outcomes (NZD, except for Employed)\** |  |  |
| Total income | 9 079 (13 047) | 12 918 (15 042) |
| Wages & salaries | 8 034 (12 900) | 11 099 (14 862) |
| Employed (N) | 34731 | 35484 |
| Benefit payments | 289 (1 103) | 338 (1 218) |
| ACC employment compensation | 112 (1 372) | 264 (1 919) |

Cells report the per-quarter mean (SD) of the outcome, unless otherwise specified
\* Only for non-work injuries

**Figure A1:**
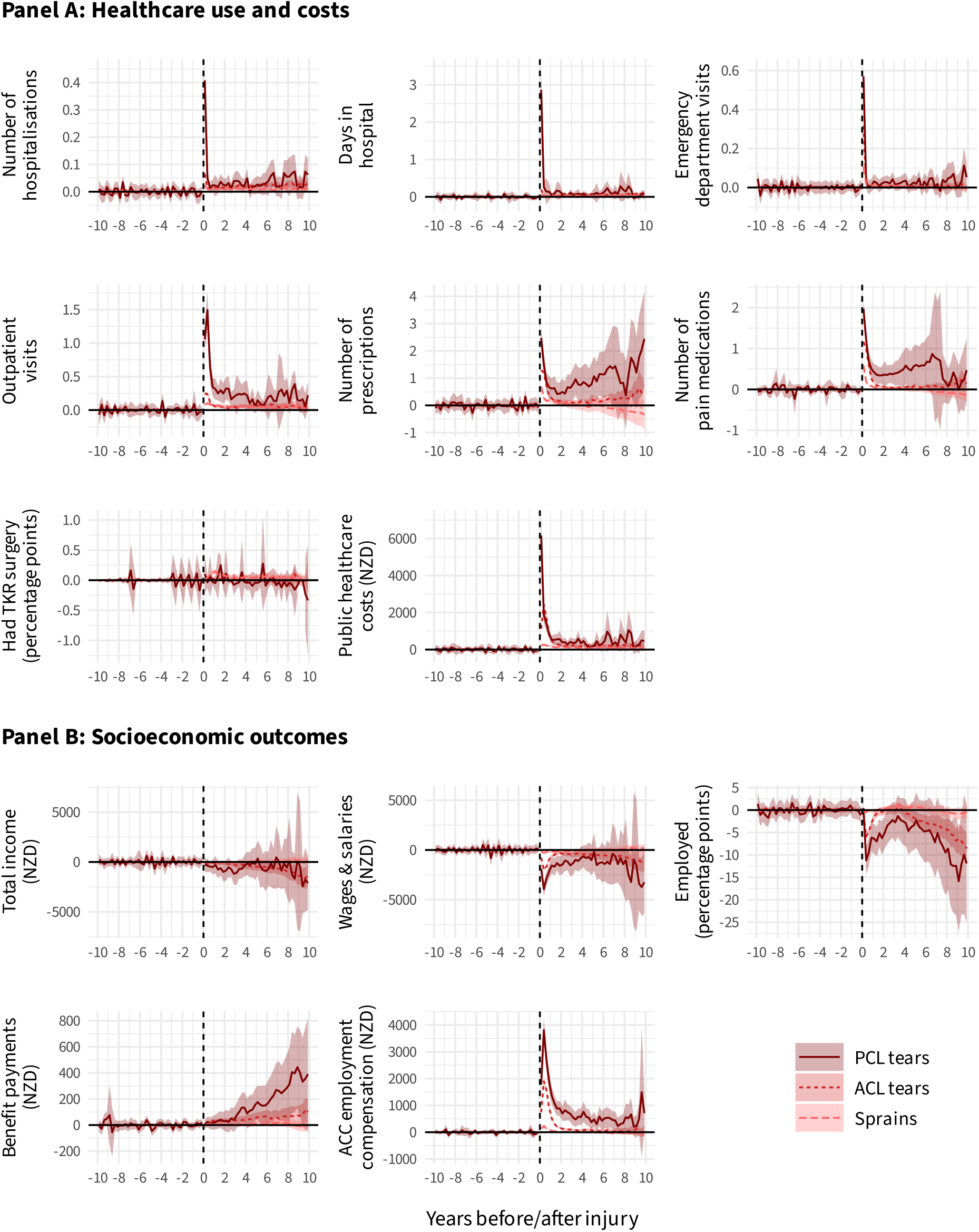
Average per-quarter effects of a cruciate ligament injury, by injury type.

**Figure A2:**
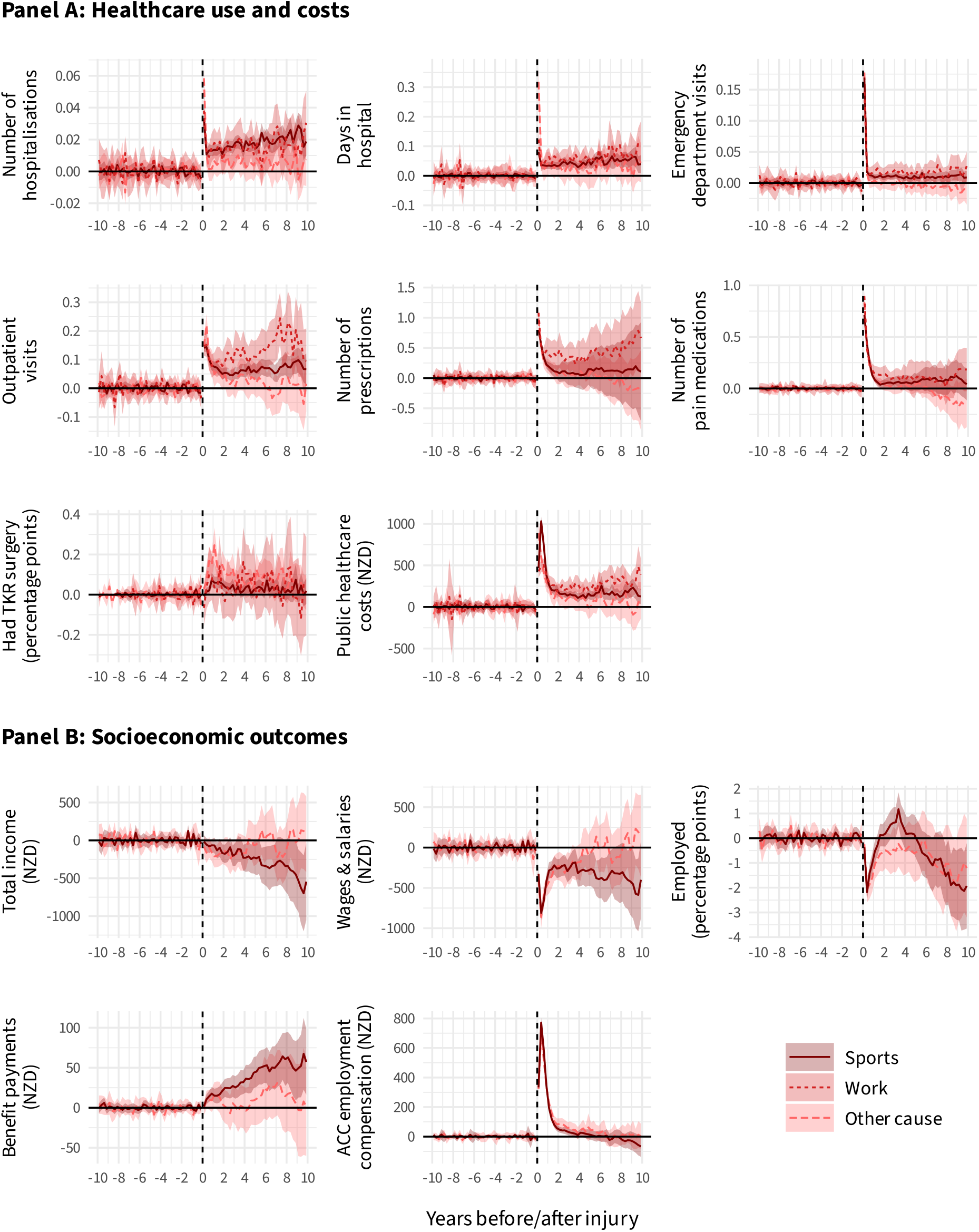
Average per-quarter effects of a cruciate ligament injury, by cause of injury.

**Figure A3:**
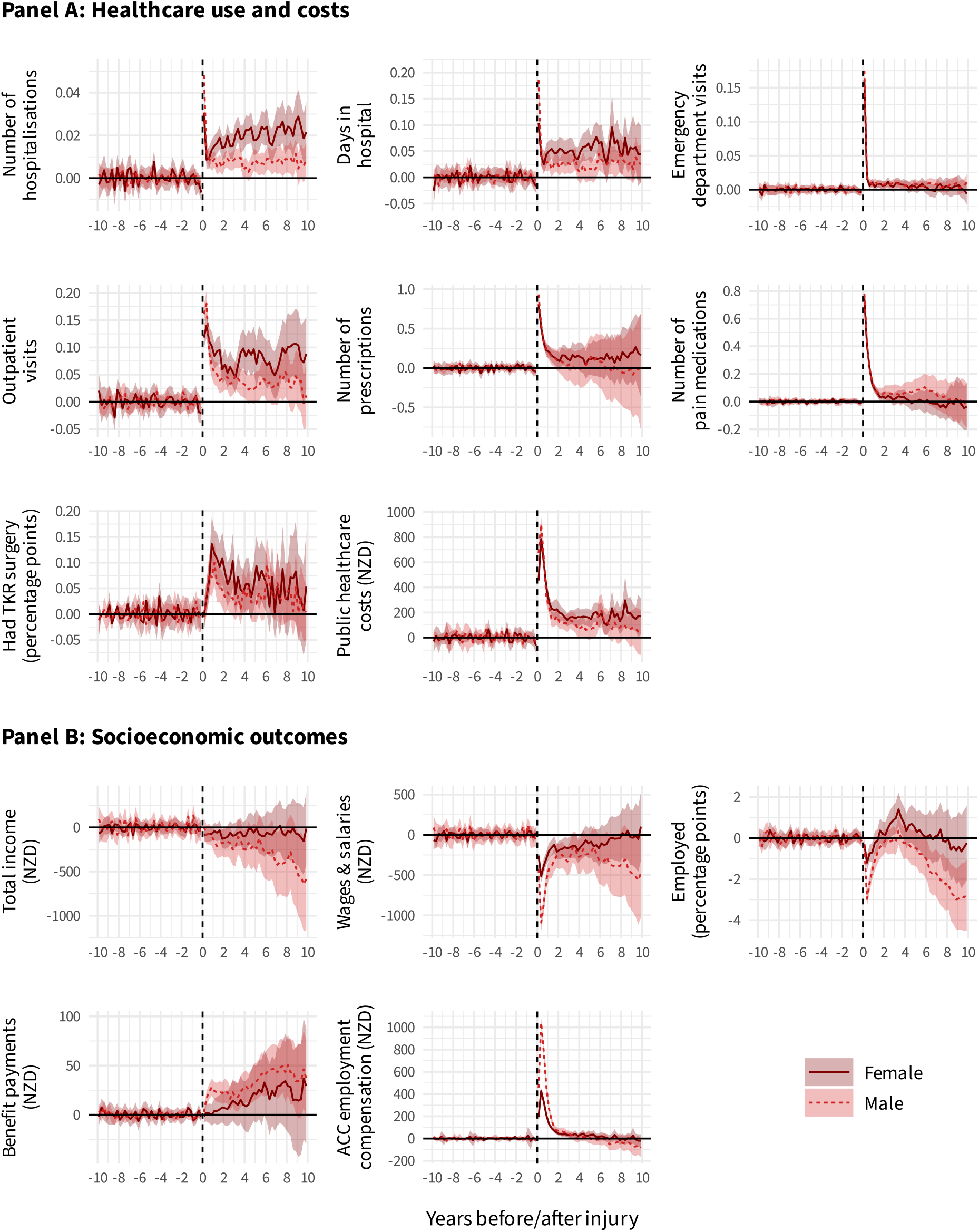
Average per-quarter effects of a cruciate ligament injury, by sex.

**Figure A4:**
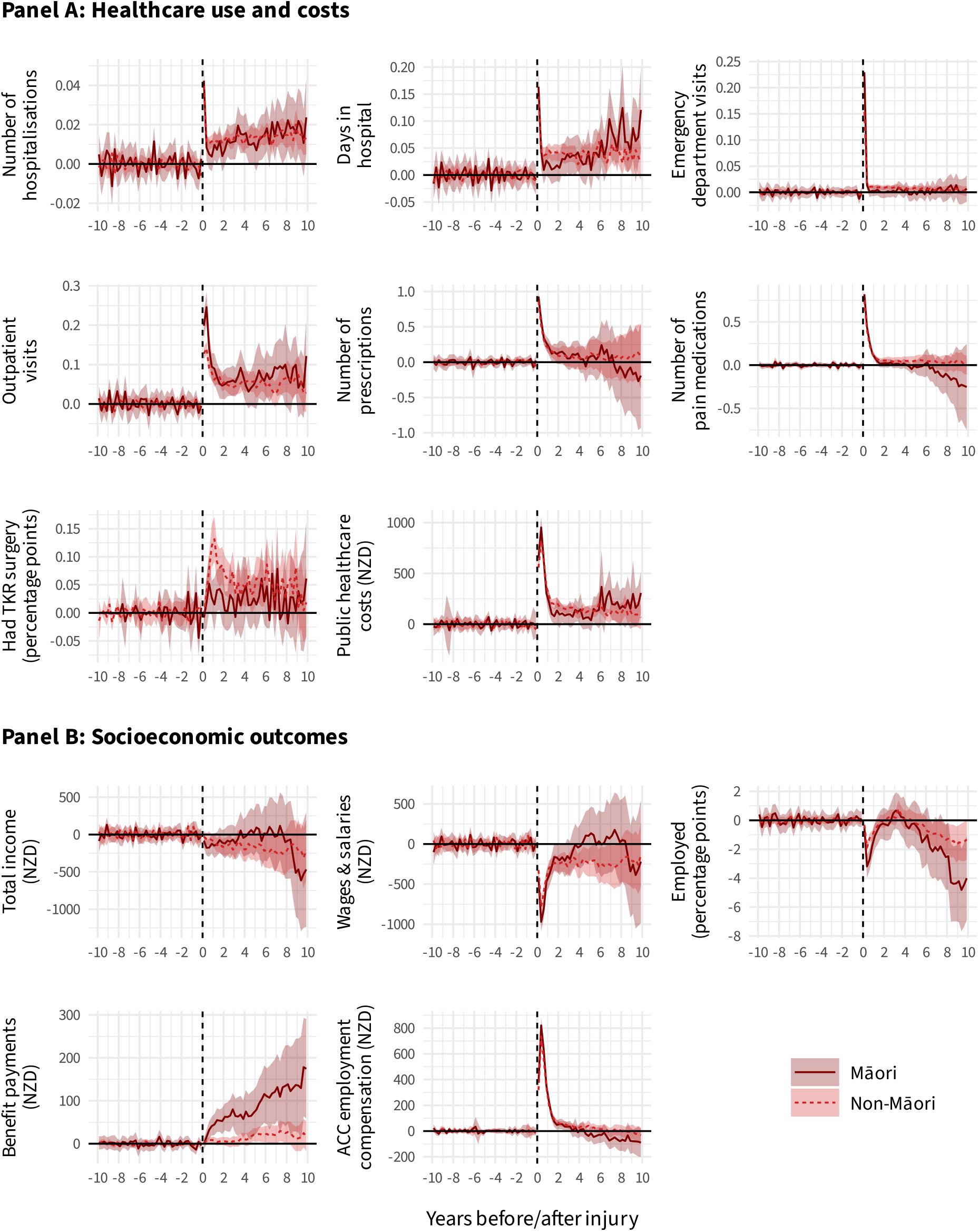
Average per-quarter effects of a cruciate ligament injury, by ethnicity.

